# Utility of genetic screening for the prediction of severe arrhythmic outcomes in mitral valve prolapse

**DOI:** 10.64898/2026.06.22.26356215

**Authors:** Rohit Jhawar, Luca Cristin, Aeron M. Small, Dwight Bibby, Lionel Tastet, Amy Rich, Francesca N. Delling

## Abstract

**Background:** Cardiomyopathy and channelopathy (CC) gene variants have been linked to sudden cardiac arrest (SCA) or death (SCD) in small, selected pedigree or post-mortem studies of arrhythmic mitral valve prolapse (MVP). However, the utility of clinical whole exome sequencing (WES) panels as a risk stratification tool in unselected MVP samples is unknown.

**Objectives:** The goal of the study was to test the utility of clinical WES panels with CC variant screening for arrhythmic risk stratification in MVP.

**Methods:** We performed research-based WES in 203 consecutive MVPs without other arrhythmic substrate. Variants were filtered for rare (<0.1%) and protein altering variants in 157 CC genes within an existing clinical panel and annotated with a clinical significance predictor. Overall frequency of CC variants was compared to a sample of general population exomes from gnomad v4.1.0. We assessed a composite severe arrhythmic outcome of SCD or frequent ectopy/ventricular tachycardia or ventricular fibrillation/SCA requiring catheter ablation or defibrillator implantation, respectively.

**Results:** CC variants were more common in MVPs compared to the general population (RR: 4.3, p < 0.01). Pathogenic/Likely Pathogenic (P/LP) variants were identified in 18 MVPs (9%; 8 CC variants among 12 genes). P/LP variants were independently associated with the composite arrhythmic outcome after adjustment for traditional imaging parameters of risk including mitral annular disjunction and bileaflet involvement (OR: 1.23 [95% CI: 1.03 – 1.47], p = 0.01). P/LP variant carriers were at greater arrhythmic risk in time-to-event analyses starting at birth (HR: 2.87 [95% CI: 1.24 – 6.62], p = 0.01).

**Conclusions:** A subset of MVPs with P/LP variants in CC genes are at higher arrhythmic risk. A clinical WES panel inclusive of CC variants may represent a valuable arrhythmic risk stratification tool in MVP beyond traditional imaging parameters.

**Clinical Perspective:** *What Is Known:*

- Cardiomyopathy/channelopathy gene variants have been linked to sudden cardiac arrest or death in small, selected pedigree or post-mortem studies of arrhythmic mitral valve prolapse (MVP).
- Imaging studies have highlighted a diffuse myopathic process in patients with arrhythmic MVP and their family members.

*What The Study Adds:*

- In consecutive MVP patients, cardiomyopathy/channelopathy genetic mutations are linked to a subclinical myopathy by strain echocardiography and a greater risk for severe ventricular arrhythmias independently of bileaflet involvement and mitral annular disjunction
- A clinical whole exome sequencing panel inclusive of cardiomyopathy/channelopathy genetic variants may represent a valuable arrhythmic risk stratification tool in MVP beyond traditional imaging parameters

Graphical Abstract:
Cardiomyopathy/Channelopathy Genetic Testing in Mitral Valve Prolapse.
Genetic samples from 203 MVPs were sequenced, and MVPs were subsequently divided based on whether they carried P/LP variants in CC genes (Top Left) or not. P/LP variants were identified in multiple genes associated with a variety of CC conditions (Bottom Left). A Kaplan-Meier curve demonstrates that P/LP variant carriers are at higher risk for the composite arrhythmic outcome beginning at birth (Right).

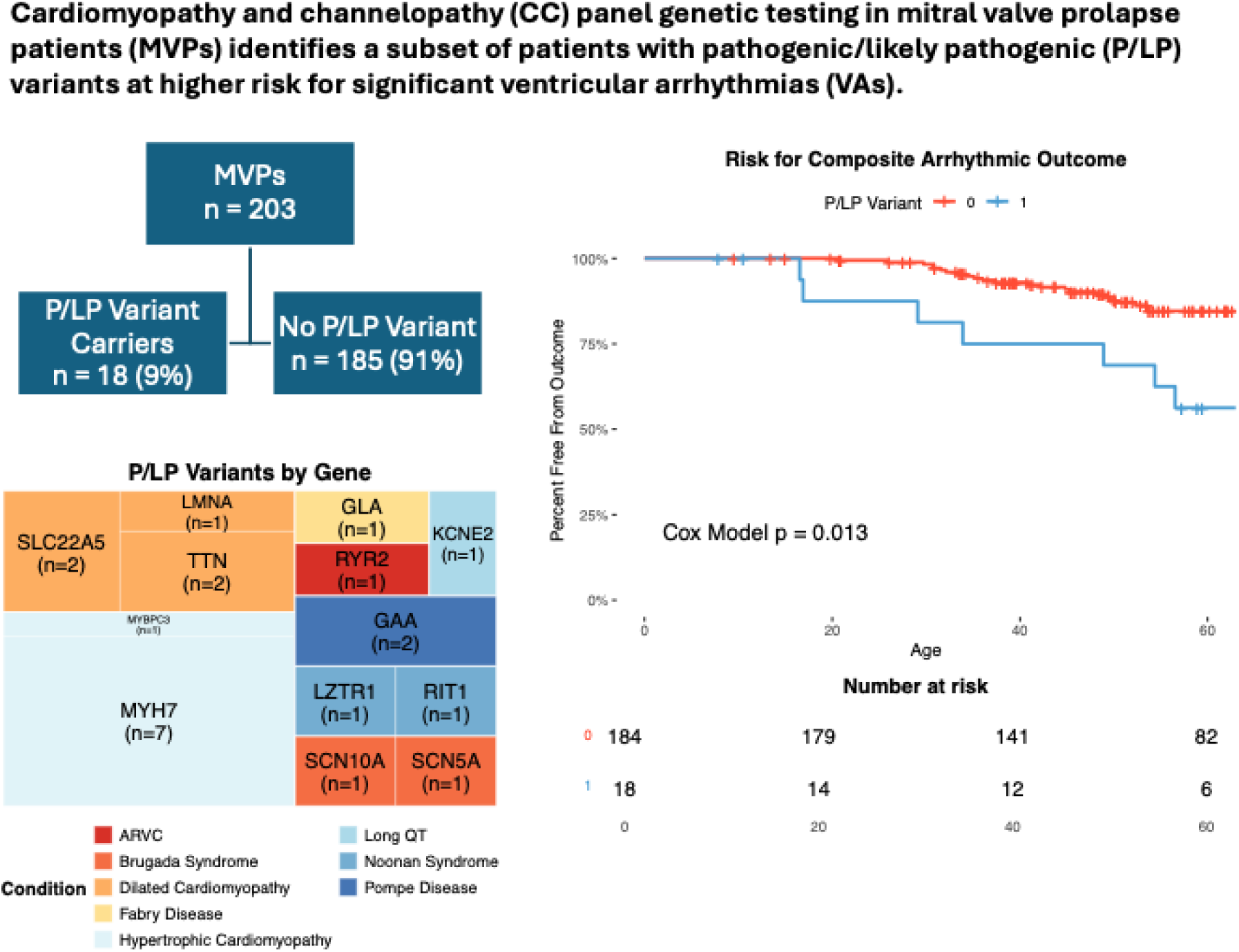

## Introduction

Mitral valve prolapse (MVP) is a valvular abnormality that affects 2-3% of the general population and is defined by echocardiography as superior displacement of the mitral leaflets into the left atrium (LA) during mid to end systole.^1, 2^ MVP is associated with severe mitral regurgitation (MR), ventricular arrythmias (VAs), and even sudden cardiac arrest (SCA) or death (SCD).^3^ Arrhythmic MVP (AMVP) has been linked to abnormal valvular-myocardial mechanics leading to papillary muscle traction by the prolapsing leaflets and focal fibrosis, particularly in those with mitral annular disjunction (MAD). The subset of MVP patients at risk for severe VAs is difficult to identify, as traditional risk factors such as bileaflet involvement, MAD, and late gadolinium enhancement (LGE) on cardiac magnetic resonance (CMR) imaging are not consistently identified.^4, 5^ As such, there is a need for additional clinical tools to risk stratify MVP patients and inform management strategies.

Recently, the presence of an underlying genetic cardiomyopathy or channelopathy (CC) has been proposed in pedigree studies of AMVP and larger genetic studies with heterogeneous presentations.^6,7^ Indeed, a diffuse myopathic process has been reported by our group and others based on speckle-tracking echocardiography (STE), diffuse fibrosis by CMR, and post-mortem data, and may be inherited prior to MVP expression.^5,6,8^ Moreover, STE-derived left ventricular (LV) mechanical dispersion (MD), defined by heterogeneous contraction/electrical dispersion, has been shown to be increased in AMVP, but also in long QT syndrome and dilated cardiomyopathy, conditions not associated with abnormal valvular-myocardial mechanics.^8–10^

Variants in cardiomyopathy genes have been identified in patients with clinically detectable myopathy with and without MVP.^11^ Testing for such variants is now widely available through clinical panels.^11^ However, CC genetic testing panels are not standardly ordered for MVP patients, especially when LV systolic function is normal or when its impairment is attributed to severe MR. Importantly, CC genetic panels are yet to be validated for arrhythmic risk stratification in MVP.

We hypothesized that testing for CC genetic variants in MVP would identify a subset of patients with pathogenic/likely pathogenic (P/LP) variants who are increased risk of significant VAs. We tested this hypothesis by analyzing whole exome sequences (WES) in consecutive MVP patients and assessing whether P/LP variants were associated with arrhythmic outcomes independently of other clinical and imaging-based arrhythmic risk factors.

## Methods

### Study Population

We obtained research-based genetic samples in 214 MVP patients at the University of California, San Francisco (UCSF) between 2017 and 2023 as part of a larger UCSF MVP Registry with available clinical, echocardiographic and ambulatory ECG monitoring.^6,12^ Genetic samples were obtained in an unbiased fashion regardless of rhythmic data. We excluded 11 MVPs due to known history of LV dysfunction (ejection fraction <50%), congenital heart disease, prior ischemic heart disease/myocardial infarction, or other phenotypically evident arrhythmic substrates (long QT, arrhythmogenic right ventricular cardiomyopathy, hypertrophic cardiomyopathy, or Brugada syndrome), with a final sample of 203 MVPs with WES. All participants signed an informed consent, and the study was approved by the UCSF Institutional Review Board.

### Variant Identification and Frequency Comparisons

WES results were filtered for high quality reads of single nucleotide polymorphisms (SNPs). SNPs were further filtered for variants of interest, defined as rare (<0.1% frequency in ExAC, a large database of sequenced human exomes),^13^ exonic, nonsynonymous variants in 157 selected CC genes (Supplementary Table 1). The list of CC genes was modeled after a widely used CC gene panel (Invitae Arrhythmia and Cardiomyopathy Comprehensive Panel with Add-On Genes, San Francisco, USA), with the intent of emulating the real-world clinical experience of ordering CC genetic testing. Deidentified variants of interest were annotated with clinical significance predictions using an artificial intelligence (AI) based algorithm (Franklin, Venlo, NL).^14,15^ Variants with P/LP predictions were considered clinically significant.

The total prevalence of both identified variants of interest and predicted P/LP variants were compared to observed frequencies in a database of 730,947 exomes from the general population (gnomAD v4.1.0, Massachusetts, USA).^16^ Frequencies were defined as the total number of variants observed in genes of interest (for the variants of interest comparison) or the total number of specific variants observed (for the P/LP variant comparison) out of the total number of exomes queried.

### Cardiac Imaging

Two-dimensional transthoracic echocardiograms were performed using commercially available ultrasound systems. All echocardiograms were reviewed by at least 2 physicians with expertise in MVP (L.C. and F.N.D.). MVP was defined as superior displacement of one or both mitral leaflets >2 mm beyond the mitral annulus in a long-axis view.^2,17^ MVP was further classified as bileaflet or monoleaflet. MR severity was assessed using a multiparametric approach as recommended by guidelines.^18^ MAD was defined as a separation between the posterior mitral leaflet/LA junction, and the basal inferolateral LV myocardium in the parasternal long-axis view.^4^ Care was taken to distinguish true MAD from pseudo-MAD, where the posterior mitral leaflet curls on itself creating a false impression of MAD.^19^ Indexed LV end-diastolic and end-systolic volumes (LVEDVI, LVESVI) were calculated as previously described.^20^

STE analysis was performed offline using commercially available software (EchoPAC, GE Healthcare) and blinded to clinical and genetic data. LV global longitudinal strain (GLS) was measured as the difference in strain value between end-diastole (i.e. 0 reference point) and end-systole. Variability analyses were previously reported for STE measurements at our institution.^21^ MD was calculated as the standard deviation of the time to peak strain across 16 LV segments. Abnormal GLS and MD were defined as greater than two standard deviations beyond average values obtained from STE analysis of 120 non-MVP controls (Supplementary Table 2), consistent with previous methods.^22^

Among 203 MVPs with WES, 134 underwent CMR. Details of CMR acquisition and image analysis are reported in the Supplemental Material.

### Primary Outcome

The primary severe arrhythmic outcome was defined^23^ as a composite of 1) frequent or complex ventricular ectopy, including ventricular tachycardia (VT), ventricular fibrillation (VF)/SCA, requiring radiofrequency catheter ablation or an implantable cardioverter defibrillator (ICD), respectively and 2) SCD. Clinical events were collected through review of electronic health records and the National Death Index. Patients with frequent (≥ 5%) premature ventricular contractions or non-sustained VT not requiring catheter ablation were not included in the severe arrhythmic outcome.

### Statistical Analyses

Continuous variables are presented as median [interquartile range (IQR)] if non-normally distributed. Continuous variables were compared between groups using Welch’s t-test. Categorical variables were presented as frequencies and percentages and were compared with Fisher’s Exact Test. Comparisons of variant frequencies were done using Poisson rate tests, as patients can carry multiple variants and individual-level variant burdens were not available from GNOMAD. Association of P/LP variants with the composite outcome was first assessed cross-sectionally with logistic regression models. Demographic/clinical covariates (age, sex, and prior valvular intervention) and imaging variables (bileaflet involvement, MAD, and MR) were included *a priori* based on previously published risk factors for AMVP.^1,4^ Results are presented as odds ratios (OR) with 95% confidence intervals (CI). Cox models were utilized for time to event analyses adjusting for sex and MV intervention (time dependent variable). The proportional hazards assumption was not violated for any covariate. Consistent with prior literature, we opted for a Cox model beginning at birth because patients with variants have carried them since birth.^24,25^ Time-to-event graphs showcase Kaplan-Meier curves, truncated when fewer than 5 participants remained at risk. P values less than 0.05 were deemed significant. All analyses were done with R version 4.5.1.

## Results

Median age of the overall sample of 203 MVPs with WES was 58 [45-72], and 111 (55%) were female. Bileaflet MVP was present in 118 (58%), MAD in 84 (41%), and moderate or greater MR in 47 (23%). Median LV ejection fraction (LVEF) was 61% [56%-66%], and prior MV intervention occurred in 55 (27%).

### Description of Variants

We identified 569 CC variants which were present in 175 MVPs (86%) and spanned 114 different genes (Supplementary Table 1). The observed frequency of CC variants in our study sample was over 4-fold greater than the frequency in the general population (RR: 4.3, p < 0.01).

When we used a previously validated AI-based algorithm to predict the clinical significance of each CC variant, we identified 222 benign/likely benign variants (39%) and 327 variants (57%) of uncertain significance. Notably, the algorithm classified 7 variants (1%) as pathogenic (P) and 13 (2%) as likely pathogenic (LP) (Table 1). We collectively referred to these as the P/LP variants and found that 18 MVPs (9%) carried a P/LP variant. The aggregate frequency of P/LP variants in our study sample was over 26-fold greater than their frequency in the general population (RR: 26.9, p < 0.01) (Supplementary Table 3).

**Table 1.**
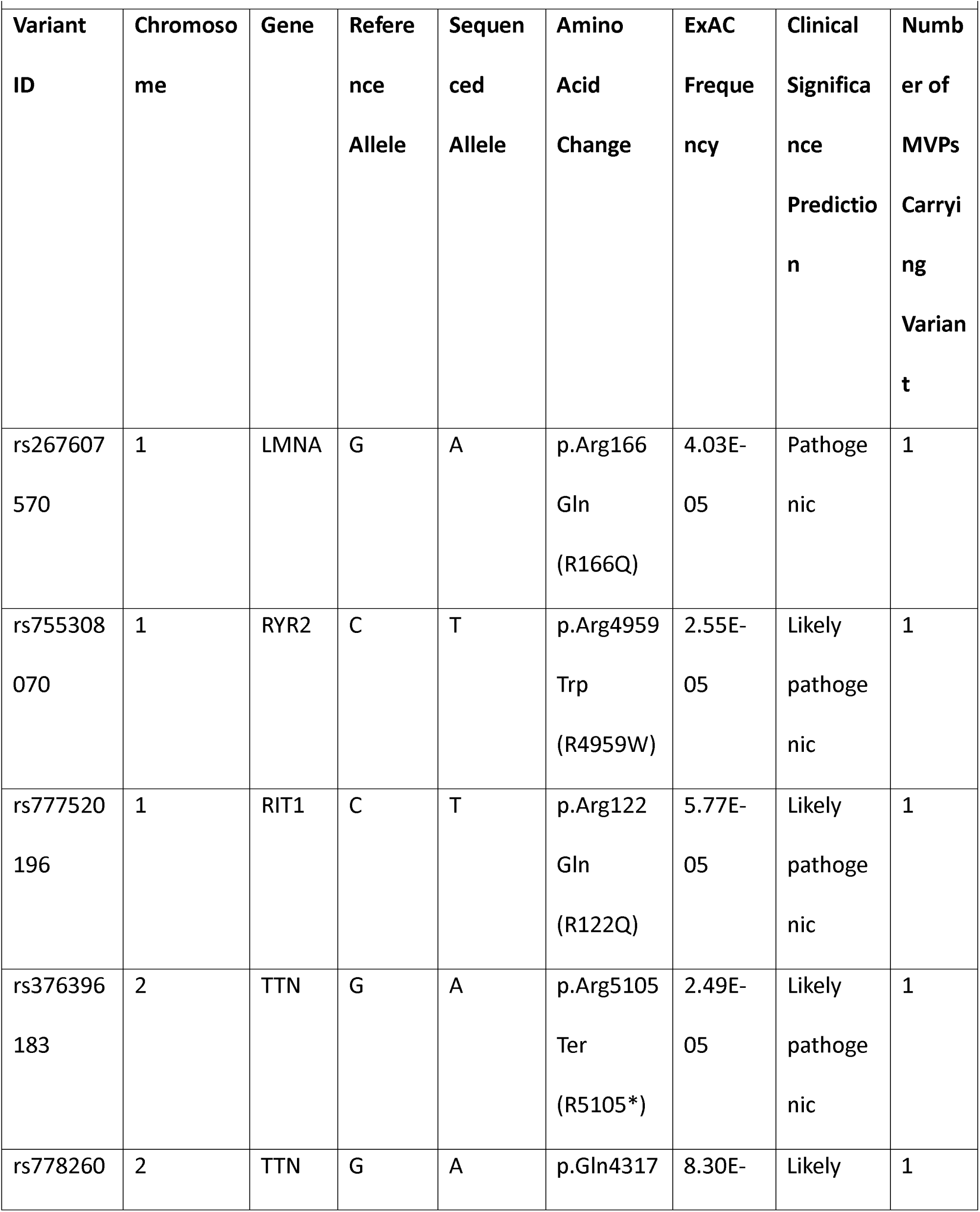
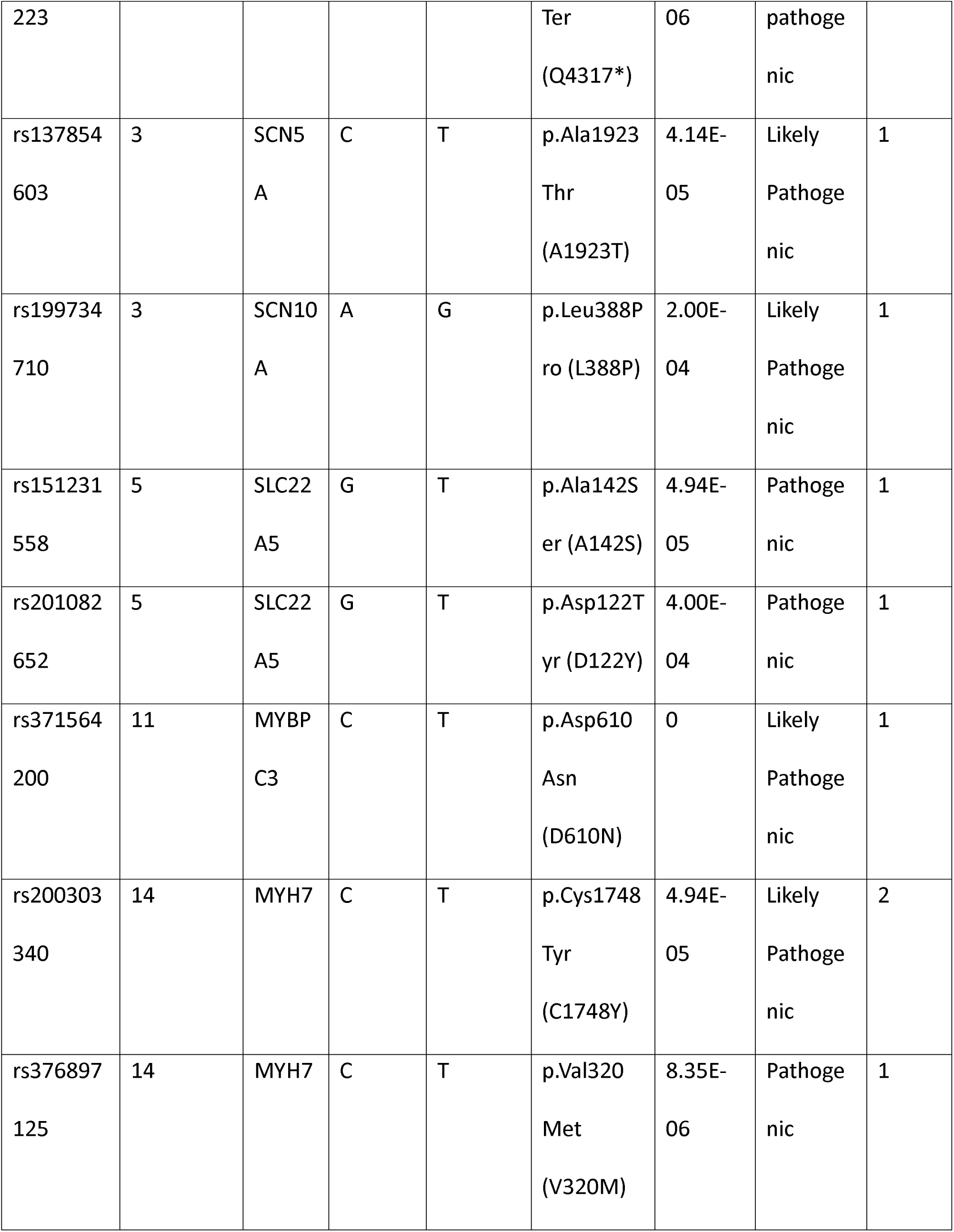
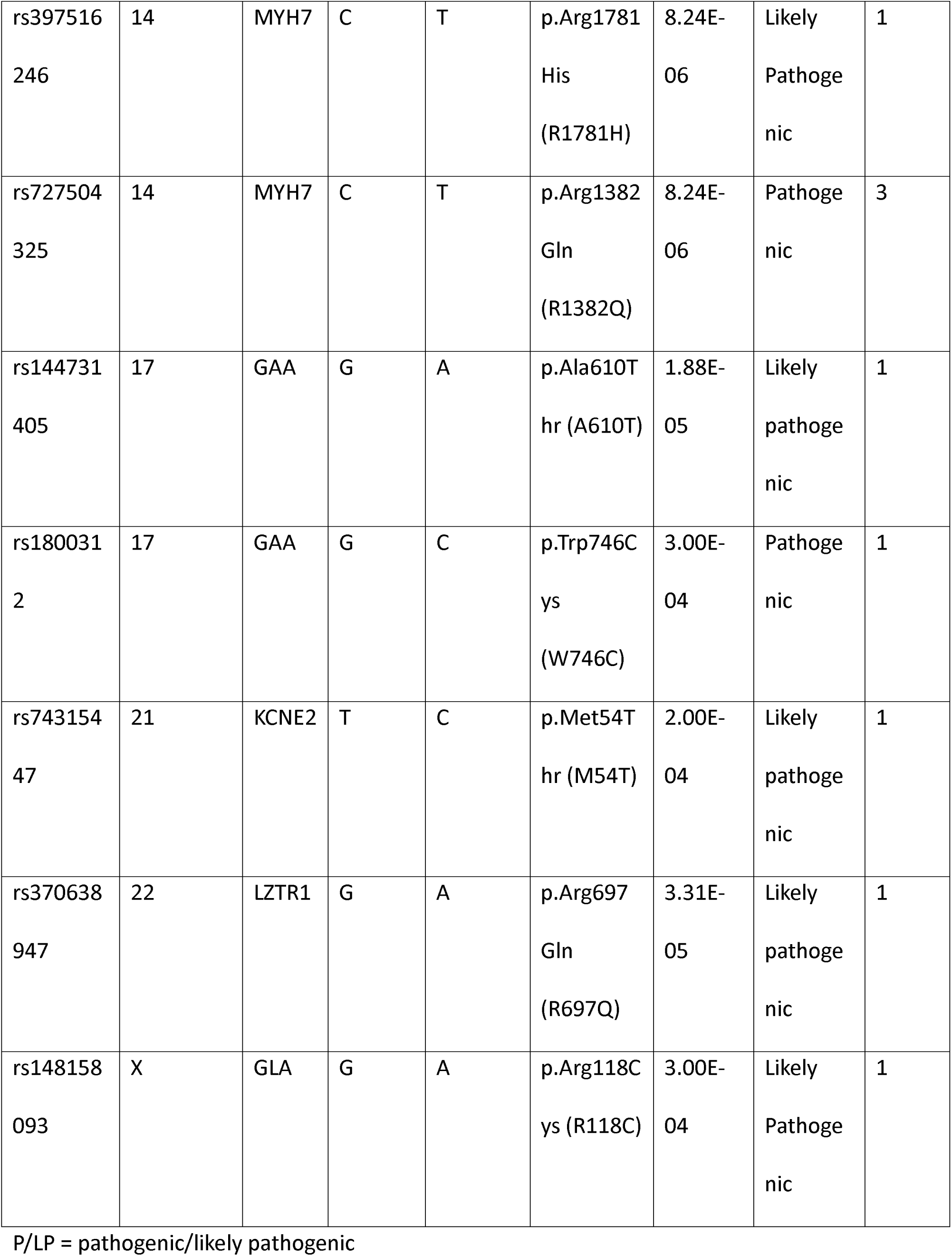
List of P/LP Variants Identified.

The P/LP variants spanned 12 different genes and 8 different CC conditions (Graphical Abstract). We reviewed each P/LP carrier’s previous cardiac imaging studies within the electronic health record to evaluate genotype-phenotype correlations. Notably, none of the P/LP variant carriers demonstrated overt phenotypic traits associated with their variant.

Specifically, none of the P/LP carriers had clear evidence of hypertrophic cardiomyopathy, arrhythmogenic cardiomyopathy, Fabry’s disease or channelopathy on cardiac imaging or electrocardiography.

### Clinical and imaging characteristics based on P/LP variant status

Our comparison of patient characteristics (Table 2) revealed no significant differences with regards to clinical or traditional arrhythmic risk imaging parameters such as MAD, MR severity, or LGE on CMR between MVPs with or without P/LP variants.^6,21^ LVEF, LV volumes and GLS were also similar. However, there was a higher proportion of MVPs with abnormal MD (2 SD above normal) in the group with compared to the one without P/LP variants. In multivariate models adjusted for age and sex, P/LP variants remained significantly associated with abnormal MD values (OR: 1.31 [95% CI: 1.07 – 1.60], p < 0.01).

**Table 2.**
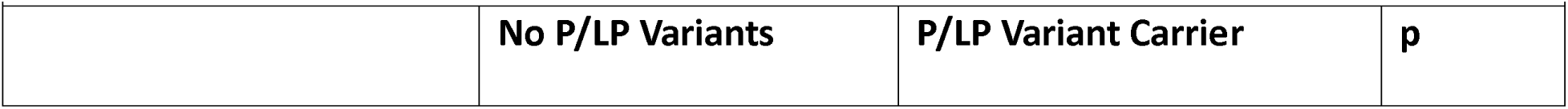
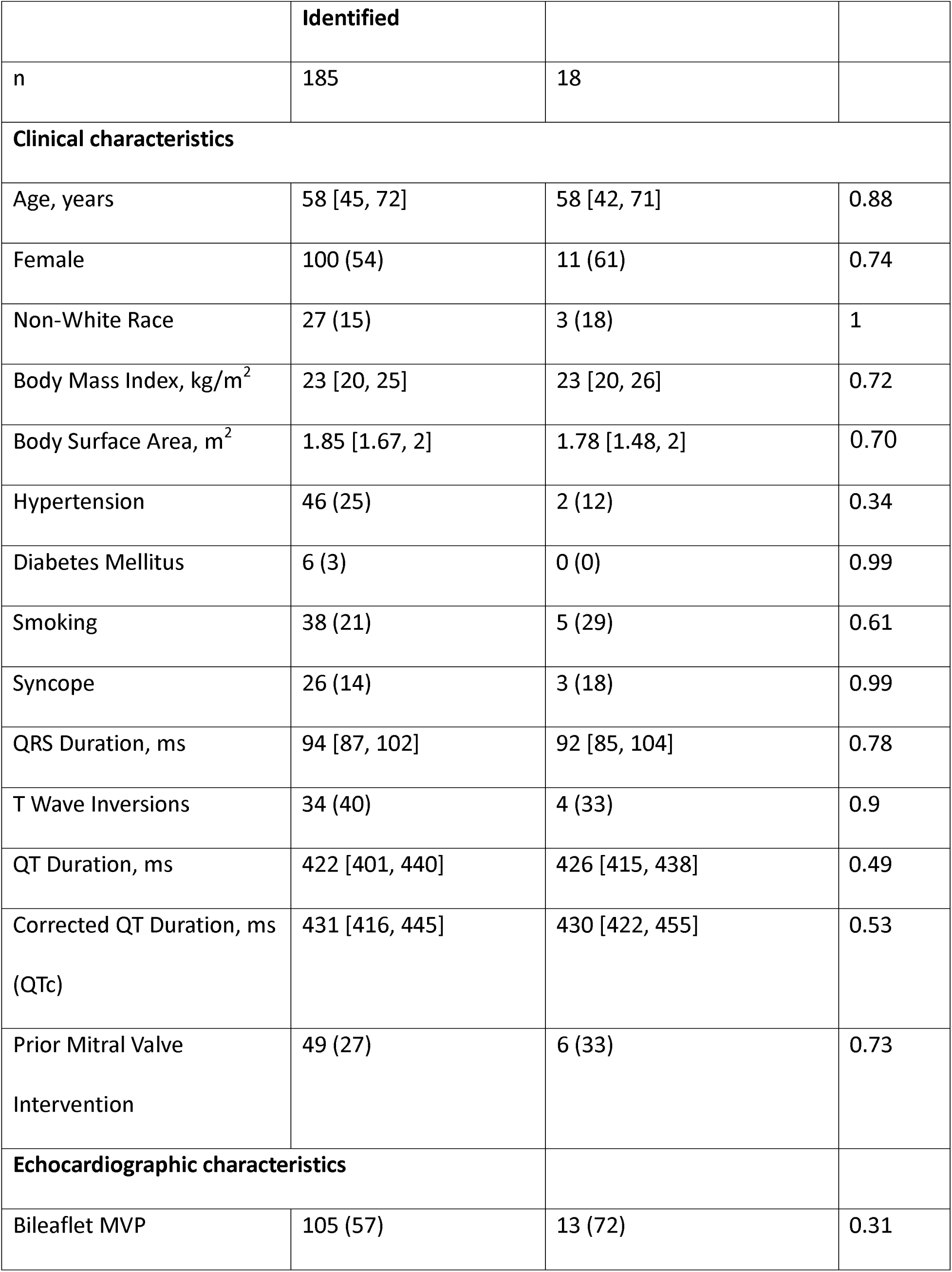
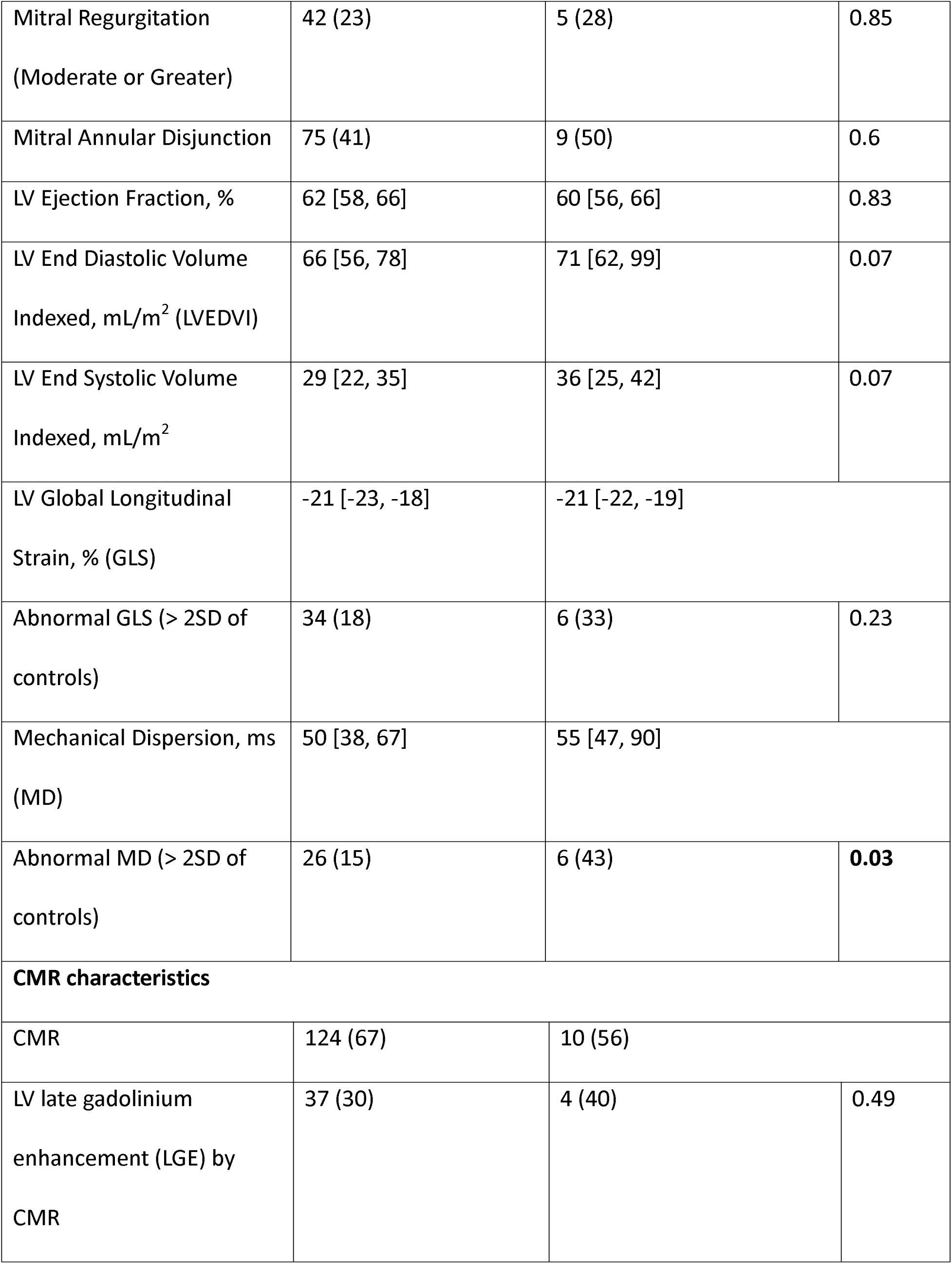
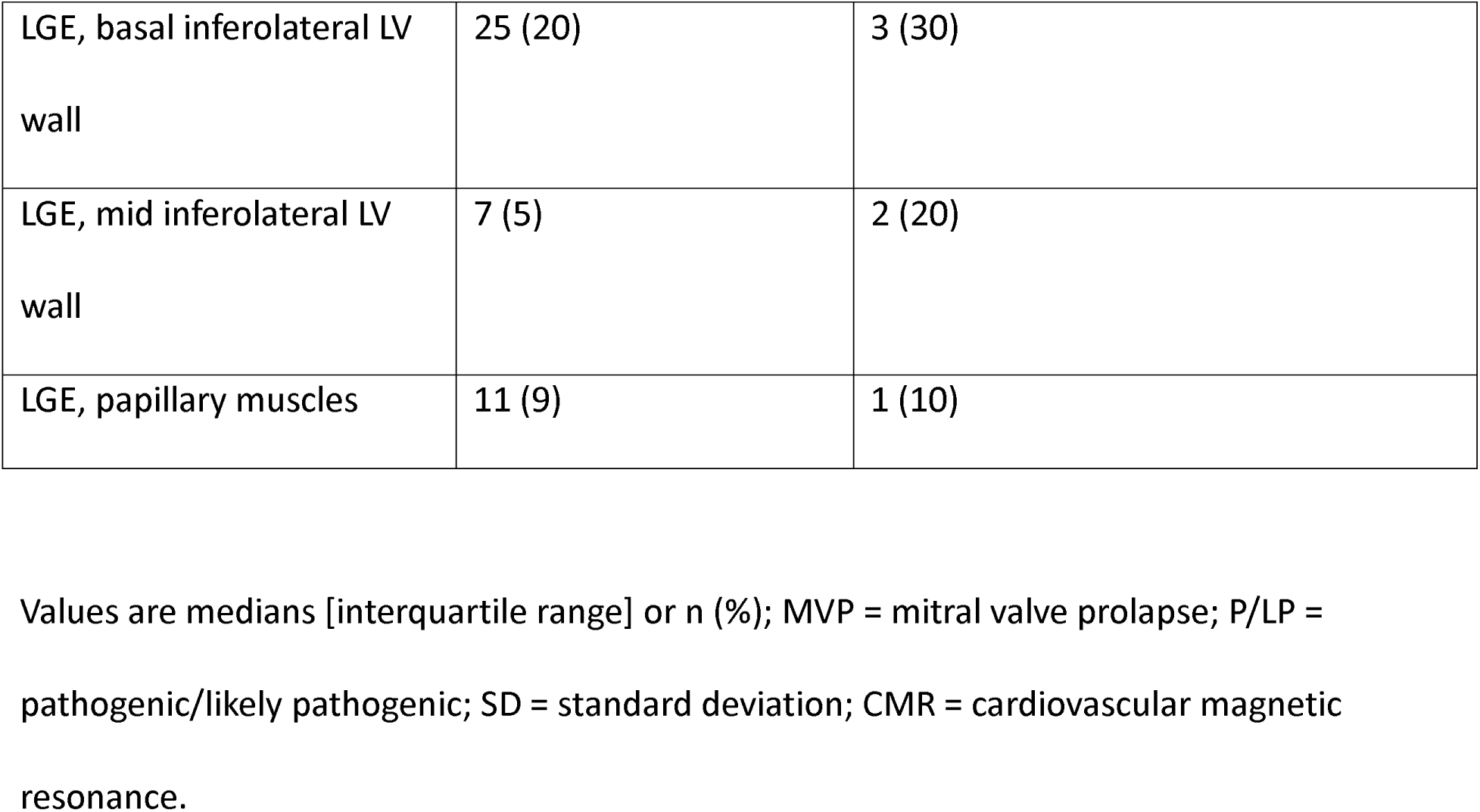
Characteristics of MVP patients, Stratified By P/LP Variants.

| Characteristics of MVP patients, Stratified By P/LP Variants |  |  |  |
| --- | --- | --- | --- |
|  | No P/LP Variants | P/LP Variant Carrier | p |
|  | **Identified** |  |  |
| n | 185 | 18 |  |
| **Clinical characteristics** |  |  |  |
| Age, years | 58 [45, 72] | 58 [42, 71] | 0.88 |
| Female | 100 (54) | 11 (61) | 0.74 |
| Non-White Race | 27 (15) | 3 (18) | 1 |
| Body Mass Index, kg/m<sup>2</sup> | 23 [20, 25] | 23 [20, 26] | 0.72 |
| Body Surface Area, m<sup>2</sup> | 1.85 [1.67, 2] | 1.78 [1.48, 2] | 0.70 |
| Hypertension | 46 (25) | 2 (12) | 0.34 |
| Diabetes Mellitus | 6 (3) | 0 (0) | 0.99 |
| Smoking | 38 (21) | 5 (29) | 0.61 |
| Syncope | 26 (14) | 3 (18) | 0.99 |
| QRS Duration, ms | 94 [87, 102] | 92 [85, 104] | 0.78 |
| T Wave Inversions | 34 (40) | 4 (33) | 0.9 |
| QT Duration, ms | 422 [401, 440] | 426 [415, 438] | 0.49 |
| Corrected QT Duration, ms<br>(QTc) | 431 [416, 445] | 430 [422, 455] | 0.53 |
| Prior Mitral Valve<br>Intervention | 49 (27) | 6 (33) | 0.73 |
| **Echocardiographic characteristics** |  |  |  |
| Bileaflet MVP | 105 (57) | 13 (72) | 0.31 |
| Mitral Regurgitation<br>(Moderate or Greater) | 42 (23) | 5 (28) | 0.85 |
| Mitral Annular Disjunction | 75 (41) | 9 (50) | 0.6 |
| LV Ejection Fraction, % | 62 [58, 66] | 60 [56, 66] | 0.83 |
| LV End Diastolic Volume<br>Indexed, mL/m<sup>2</sup> (LVEDVI) | 66 [56, 78] | 71 [62, 99] | 0.07 |
| LV End Systolic Volume<br>Indexed, mL/m<sup>2</sup> | 29 [22, 35] | 36 [25, 42] | 0.07 |
| LV Global Longitudinal<br>Strain, % (GLS) | -21 [-23, -18] | -21 [-22, -19] |  |
| Abnormal GLS (> 2SD of<br>controls) | 34 (18) | 6 (33) | 0.23 |
| Mechanical Dispersion, ms<br>(MD) | 50 [38, 67] | 55 [47, 90] |  |
| Abnormal MD (> 2SD of<br>controls) | 26 (15) | 6 (43) | 0.03 |
| **CMR characteristics** |  |  |  |
| CMR | 124 (67) | 10 (56) |  |
| LV late gadolinium<br>enhancement (LGE) by<br>CMR | 37 (30) | 4 (40) | 0.49 |

|  |  |  |
| --- | --- | --- |
| LGE, basal inferolateral LV wall | 25 (20) | 3 (30) |
| LGE, mid inferolateral LV wall | 7 (5) | 2 (20) |
| LGE, papillary muscles | 11 (9) | 1 (10) |

Values are medians [interquartile range] or n (%); MVP = mitral valve prolapse; P/LP =
pathogenic/likely pathogenic; SD = standard deviation; CMR = cardiovascular magnetic
resonance.

### Association of variants with arrhythmic outcomes and subclinical myopathy

In our sample of consecutive MVP patients, we sought to investigate whether P/LP variants were associated with the composite arrhythmic outcome despite lack of overt CC phenotypes. In total, 35 MVPs (17%) experienced the composite outcome, of which 1 experienced SCD, 20 had a SCA/ICD, and 14 had frequent ectopy or VT requiring either a catheter ablation or ICD. The composite arrhythmic outcome occurred significantly more often in P/LP variant carriers compared to non-carriers (39% vs 15%, p = 0.01). P/LP variants remained a significant predictor of the composite outcome in models adjusted for demographic/clinical variables (OR: 1.25 [95% CI: 1.04 – 1.49], p = 0.02) and imaging variables (OR: 1.23 [95% CI: 1.03 – 1.47], p = 0.02) (Table 3). Consistent with prior literature,^26,27^ bileaflet prolapse was also a significant predictor of the composite outcome.

**Table 3.** Association of P/LP Variants With the Composite Arrhythmic Outcome. Results are Odds Ratio (OR) with 95% CI. Two separate multivariate models were run, with one including demographic/clinical Variables (age, sex, prior valvular intervention) and the other including imaging variables (bileaflet prolapse, MAD, mitral regurgitation). P/LP = pathogenic/likely pathogenic; MAD = mitral annular disjunction.

| **Association of P/LP Variants With the Composite Arrhythmic Outcome** |  |  |
| --- | --- | --- |
|  | **Univariate Analysis** | **Multivariate Analysis** |

| Predictor | OR (95% CI) | p Value | OR (95% CI) | p Value |
| --- | --- | --- | --- | --- |
| **Demographic/Clinical Variables** |  |  |  |  |
| P/LP Variant | 1.27 (1.06 - 1.52) | **0.01** | 1.25 (1.04 - 1.49) | **0.02** |
| Age | 1 (0.99 - 1) | 0.38 | 1.0 (0.99 - 1.0) | 0.22 |
| Sex | 1.06 (0.95 - 1.18) | 0.29 | 1.07 (0.97 - 1.19) | 0.18 |
| Prior Valvular Intervention | 1.09 (0.97 - 1.23) | 0.14 | 1.12 (1 - 1.27) | 0.06 |
| **Imaging Variables** |  |  |  |  |
| P/LP Variant | 1.27 (1.06 - 1.52) | **0.01** | 1.23 (1.03 - 1.47) | **0.02** |
| Bileaflet | 1.24 (1.12 - 1.37) | **<0.01** | 1.23 (1.11 - 1.36) | **<0.01** |
| MAD | 1.01 (0.91 - 1.12) | 0.85 | 1.01 (0.91 - 1.12) | 0.58 |
| Mitral Regurgitation (Moderate or greater) | 1 (0.88 - 1.13) | 0.96 | 0.98 (0.87 - 1.1) | 0.41 |

In an adjusted Cox proportional hazards model beginning at birth, P/LP variant carriers were at significantly greater risk for the composite arrhythmic outcome (HR: 2.87 [95% CI: 1.24 – 6.62], p = 0.01) (Graphical Abstract). The median age at analysis was 62 years [58 – 66].

## Discussion

Our study evaluated the clinical utility of genetic panel testing for CC variants in a sample of consecutive MVP patients with available research-based genetic samples, rhythmic data, and imaging assessment. Because study data was obtained in all-comers and independently of arrhythmic presentation, our investigation provides an unbiased assessment of the prevalence and clinical significance of CC genetic variants in MVP. Our key findings are as follows: 1) MVPs frequently carry CC variants, with a prevalence significantly higher than the general population; 2) MVPs carrying P/LP variants for CC genes are at higher risk of developing a composite outcome of frequent or complex ventricular ectopy, including VT and VF/SCA, requiring catheter ablation or implantation of an ICD in a time-to-event analysis; 3) MVPs carrying P/LP variants for CC genes are also more likely to have abnormal LV mechanical/electrical dispersion, linking genetics to the presence of a subclinical myopathy.^5,21,28,29^

A prior meta-analysis of six genome-wide association studies highlighted for the first time the role of cardiomyopathy genes in MVP, leading to the creation of a polygenic risk score able to predict development of MVP better than age, sex, and clinical risk factors.^30^ Similarly, we found a higher number of CC variants overall among MVP patients compared to a sample of the general population when using a commercially available targeted exome sequencing panel. This may further support increased yield from clinical genetic testing in the MVP population.

Regardless of prediction of arrhythmic outcomes, knowledge of cardiomyopathy genes may be useful in predicting adverse remodeling despite MV repair or in deciding about earlier intervention for significant MR.

The previously cited GWAS highlighted the role of cardiomyopathy genes in MVP but did not link these genes with arrhythmic complications. Other smaller studies have suggested arrhythmic MVPs carry more P/LP variants in CC genes overall compared to other patients with severe VAs or the general population.^7,31,32^ However, these studies have included mostly case reports of SCA or post-mortem MVPs with arrhythmic death. In our investigation we demonstrate the utility of screening for CC variants with a readily available clinical panel in any MVP patient regardless of clinical presentation and prior to the development of severe arrhythmic outcomes. To the best of our knowledge, we are the first to report that P/LP variants in CC genes in MVP may have prognostic value in predicting significant VAs. This was made possible by the availability of unselected research samples from our UCSF MVP Registry, whereas typically clinical genetic panels are selectively used only in patients that have already presented with a severe arrhythmic outcome. In the future, clinical genetic panels may be used to screen MVP patients that may benefit from closer follow-up prior to the development of significant VAs.

Interestingly, P/LP variants in CC genes were identified in AMVPs that didn’t necessarily exhibit the clinical phenotype (on imaging or ECG) of a particular cardiomyopathy or channelopathy. Whether CC genes create an electrical substrate for the development of VAs that precedes the development of structural abnormalities of the myocardium needs to be further investigated in both MVP and non-MVP samples. Moreover, it is still unclear whether an underlying arrhythmogenic genetic substrate that weakens myocyte coupling may be exacerbated by traction exerted by the prolapsing leaflets on the myocardium, the “2-hit” model^31^, with MAD augmenting such abnormal valvular-myocardial mechanics. This scenario would explain why most patients with uncomplicated myxomatous MVP do not develop potentially life-threatening VAs.

Current approaches to arrhythmic risk stratification in MVPs largely rely on imaging findings like bileaflet prolapse and MAD.^4^ Our results suggest that P/LP variants in CC genes can confer increased arrhythmic risk in MVP independently of such traditional imaging parameters. This fits with a growing body of literature indicating significant residual risk for VAs in MVPs even in the absence of bileaflet involvement or MAD.^12,33^ The presence of an underlying myopathy in MVPs may explain this residual risk.^7,28,34,35^ We have demonstrated that in the familial setting family members of MVP cases may have global abnormal strain parameters despite having normal LVEF and no valve abnormality.^6^ Our results add to this hypothesis, as MVPs with P/LP variants had more abnormal MD values.^36,37^ Further mechanistic studies are necessary to unravel the causal relationships between genetics, myocardial function, valvular hemodynamics, and VAs in MVPs.

The goal of our study was to mimic the clinical experience of ordering CC gene panel tests on MVP patients. Further research, including replication studies, cost-effectiveness estimates, and prospective trials, may build on the initial positive results presented here to establish the clinical validity of panel genetic testing for arrhythmic risk stratification in MVP.

### Limitations

This analysis was conducted at a single center using a single sequencing platform and variant effect prediction algorithm, limiting external validity and generalizability. Outcomes were adjudicated based on clinical records and were not prospectively measured. A certified genetic counselor was not available to validate the AI algorithm’s predictions.

### Conclusion

Variants in cardiomyopathy and channelopathy genes are more common in MVP compared to the general population. MVPs with P/LP variants are at increased risk for significant ventricular arrhythmias including sudden cardiac arrest and death. A clinical genetic panel inclusive of CC variants may represent a valuable arrhythmic risk stratification tool in MVP beyond traditional imaging parameters.

## Data Availability

All data produced in the present study are available upon reasonable request to the authors. All data produced in the present work are contained in the manuscript.

## Acknowledgments

None.

---

**Supplemental Table 1.** CC Genes Tested.

| **CC Genes Tested** |  |  |  |
| --- | --- | --- | --- |
| **Gene** | **MVP Variant Counts** | **General Population**<br>**Variant Count** | **p-Value** |
| TTN | 133 | 88105 | <0.01 |
| AKAP9 | 19 | 12013 | <0.01 |
| ALMS1 | 17 | 9918 | <0.01 |
| FLNC | 16 | 9647 | <0.01 |
| SCN10A | 16 | 7044 | <0.01 |
| DMD | 14 | 13756 | <0.01 |
| DSP | 12 | 6580 | <0.01 |
| MYH7 | 11 | 6613 | <0.01 |
| RYR2 | 10 | 25318 | 0.17 |
| LDB3 | 10 | 3755 | <0.01 |
| AGL | 9 | 5975 | <0.01 |
| CTNNA3 | 9 | 3836 | <0.01 |
| MYBPC3 | 8 | 9291 | 0.01 |
| HCN4 | 8 | 3993 | <0.01 |
| PRDM16 | 7 | 5409 | <0.01 |
| MYH6 | 7 | 8448 | 0.01 |
| SLC22A5 | 7 | 2265 | <0.01 |
| CACNB2 | 7 | 6139 | <0.01 |
| A2ML1 | 7 | 6400 | <0.01 |
| CACNA1C | 7 | 13415 | 0.08 |
| LAMA4 | 6 | 7451 | 0.02 |
| MYPN | 6 | 4725 | <0.01 |
| BAG3 | 6 | 1688 | <0.01 |
| TRPM4 | 5 | 6432 | 0.04 |
| ANK2 | 5 | 11802 | 0.23 |
| SDHA | 5 | 3593 | <0.01 |
| PLEKHM2 | 5 | 6773 | 0.04 |
| KCNA5 | 5 | 1451 | <0.01 |
| ACADVL | 5 | 4089 | 0.01 |
| ALPK3 | 5 | 5099 | 0.02 |
| SCN5A | 5 | 7702 | 0.07 |
| ACTN2 | 5 | 4191 | 0.01 |
| TXNRD2 | 5 | 5208 | 0.02 |
| TRDN | 4 | 10576 | 0.34 |
| LZTR1 | 4 | 6227 | 0.10 |
| RBM20 | 4 | 4446 | 0.04 |
| TMEM43 | 4 | 2673 | 0.01 |
| CACNA1D | 4 | 10169 | 0.31 |
| PRKAG2 | 4 | 3899 | 0.03 |
| KCNJ2 | 4 | 481 | <0.01 |
| PKP2 | 4 | 3797 | 0.02 |
| GAA | 4 | 5002 | 0.05 |
| TMEM70 | 3 | 1319 | 0.01 |
| VCL | 3 | 3657 | 0.08 |
| NEBL | 3 | 7152 | 0.32 |
| DTNA | 3 | 3893 | 0.1 |
| ABCC9 | 3 | 6716 | 0.29 |
| MYOM1 | 3 | 7819 | 0.37 |
| SOS2 | 3 | 6929 | 0.3 |
| LRRC10 | 3 | 521 | <0.01 |
| ELAC2 | 3 | 4755 | 0.15 |
| DSC2 | 3 | 3252 | 0.06 |
| KRAS | 3 | 961 | <0.01 |
| EYA4 | 3 | 3340 | 0.07 |
| CASQ2 | 3 | 2751 | 0.04 |
| MED12 | 3 | 5978 | 0.23 |
| NEXN | 2 | 2596 | 0.16 |
| SPRED1 | 2 | 1570 | 0.07 |
| DSG2 | 2 | 3720 | 0.28 |
| MYOZ2 | 2 | 825 | 0.02 |
| PTPN11 | 2 | 2749 | 0.18 |
| NKX2-5 | 2 | 1891 | 0.1 |
| CDH2 | 2 | 4195 | 0.33 |
| MYLK2 | 2 | 2433 | 0.15 |
| GATA6 | 2 | 2650 | 0.17 |
| TTR | 2 | 539 | 0.01 |
| GATA5 | 2 | 3215 | 0.23 |
| MYLK3 | 2 | 3280 | 0.23 |
| KIF20A | 2 | 2734 | 0.18 |
| PDLIM3 | 2 | 1965 | 0.10 |
| CPT2 | 2 | 2321 | 0.14 |
| FKTN | 1 | 3018 | 0.57 |
| SGCD | 1 | 2117 | 0.45 |
| BRAF | 1 | 5371 | 0.78 |
| SNTA1 | 1 | 2481 | 0.5 |
| MAP2K2 | 1 | 3580 | 0.63 |
| SCN4B | 1 | 1093 | 0.26 |
| RIT1 | 1 | 1022 | 0.25 |
| SCN1B | 1 | 1773 | 0.39 |
| LAMP2 | 1 | 1670 | 0.37 |
| NF1 | 1 | 10842 | 0.95 |
| PPA2 | 1 | 3033 | 0.57 |
| TPM1 | 1 | 3441 | 0.62 |
| KCNE2 | 1 | 237 | 0.06 |
| RRAS | 1 | 1736 | 0.38 |
| ANKRD1 | 1 | 1776 | 0.39 |
| KLF10 | 1 | 1506 | 0.34 |
| CSRP3 | 1 | 944 | 0.23 |
| CACNA2D1 | 1 | 9068 | 0.92 |
| TCAP | 1 | 571 | 0.15 |
| SOS1 | 1 | 5889 | 0.81 |
| MYL3 | 1 | 918 | 0.23 |
| NPPA | 1 | 634 | 0.16 |
| DOLK | 1 | 811 | 0.20 |
| PCCA | 1 | 6308 | 0.83 |
| GLA | 1 | 1335 | 0.31 |
| CTF1 | 1 | 1858 | 0.40 |
| SLMAP | 1 | 5061 | 0.76 |
| RANGRF | 1 | 753 | 0.19 |
| CALR3 | 1 | 1934 | 0.42 |
| RASA1 | 1 | 5257 | 0.77 |
| HAND1 | 1 | 796 | 0.2 |
| TNNT2 | 1 | 2646 | 0.52 |
| RASA2 | 1 | 5389 | 0.78 |
| TMPO | 1 | 2834 | 0.55 |
| MYL4 | 1 | 1024 | 0.25 |
| EMD | 1 | 1149 | 0.27 |
| LMNA | 1 | 3889 | 0.66 |
| RAF1 | 1 | 3110 | 0.58 |
| MTO1 | 1 | 3398 | 0.61 |
| KCNJ5 | 1 | 837 | 0.21 |
| ILK | 1 | 1969 | 0.42 |
| JPH2 | 1 | 3049 | 0.57 |
| ACTC1 | 0 | 577247 | <0.01 |
| CALM1 | 0 |  |  |
| CALM2 | 0 |  |  |
| CALM3 | 0 |  |  |
| CBL | 0 |  |  |
| CRYAB | 0 |  |  |
| DES | 0 |  |  |
| DNAJC19 | 0 |  |  |
| FHL1 | 0 |  |  |
| FKRP | 0 |  |  |
| GATA4 | 0 |  |  |
| GJA5 | 0 |  |  |
| HRAS | 0 |  |  |
| JUP | 0 |  |  |
| KCNE1 | 0 |  |  |
| KCNH2 | 0 |  |  |
| KCNQ1 | 0 |  |  |
| MAP2K1 | 0 |  |  |
| MRAS | 0 |  |  |
| MYL2 | 0 |  |  |
| NRAS | 0 |  |  |
| PCCB | 0 |  |  |
| PLN | 0 |  |  |
| PPCS | 0 |  |  |
| PPP1CB | 0 |  |  |
| SHOC2 | 0 |  |  |
| TAZ | 0 |  |  |
| TBX20 | 0 |  |  |
| TNNC1 | 0 |  |  |
| TNNI3 | 0 |  |  |
| TNNI3K | 0 |  |  |
| CAV3 | 0 |  |  |
| CHRM2 | 0 |  |  |
| FHL2 | 0 |  |  |
| GATAD1 | 0 |  |  |
| GPD1L | 0 |  |  |
| KCND3 | 0 |  |  |
| KCNE3 | 0 |  |  |
| KCNE5 | 0 |  |  |
| KCNJ8 | 0 |  |  |
| KCNK3 | 0 |  |  |
| MAP3K8 | 0 |  |  |
| SCN2B | 0 |  |  |
| SCN3B | 0 |  |  |

List of genes tested for deleterious variants. MVP variant counts are raw counts out of 203
MVPs tested, while general population variant counts are raw counts out of 730,947 exomes
queried from gnomad v4.1.0. CC = cardiomyopathy/channelopathy; MVP = mitral valve prolapse.

**Supplemental Table 2.** Speckle Tracking Echocardiography from 120 Non MVP Controls.

| **Speckle Tracking Echocardiography from 120 Non MVP Controls** |  |  |
| --- | --- | --- |
| **Age, years** | **Global Longitudinal Strain, %** | **Mechanical Dispersion, ms** |
| 46 (24) | -22 (3) | 42 (15) |

Values presented as mean (standard deviation). MVP = mitral valve prolapse.

**Supplemental Table 3.** CC Variants Identified.

| CC Variants Identified |  |  |  |  |
| --- | --- | --- | --- | --- |
| Variant ID | Count Identified in MVPs | Frequency in MVPs | Frequency in General Population | p-Value |
| rs267607570 | 1 | 0.01 | 3.41E-05 | <0.01 |
| rs755308070 | 1 | 0.01 | 2.47E-05 | 0.01 |
| rs777520196 | 1 | 0.01 | 1.71E-05 | <0.01 |
| rs376396183 | 1 | 0.01 | 1.45E-05 | <0.01 |
| rs778260223 | 1 | 0.01 | 2.74E-06 | <0.01 |
| rs137854603 | 1 | 0.01 | 2.53E-05 | <0.01 |
| rs199734710 | 1 | 0.01 | 9.99E-05 | 0.02 |
| rs151231558 | 1 | 0.01 | 2.68E-04 | 0.05 |
| rs201082652 | 1 | 0.01 | 6.67E-04 | 0.13 |
| rs371564200 | 1 | 0.01 | 1.24E-05 | <0.01 |
| rs200303340 | 2 | 0.01 | 3.83E-05 | <0.01 |
| rs376897125 | 1 | 0.01 | 1.37E-06 | <0.01 |
| rs397516246 | 1 | 0.01 | 1.57E-05 | <0.01 |
| rs727504325 | 3 | 0.02 | 8.90E-06 | <0.01 |
| rs144731405 | 1 | 0.01 | 3.70E-05 | <0.01 |
| rs1800312 | 1 | 0.01 | 4.91E-04 | 0.1 |
| rs74315447 | 1 | 0.01 | 1.40E-04 | 0.03 |
| rs370638947 | 1 | 0.01 | 3.15E-05 | <0.01 |
| rs148158093 | 1 | 0.01 | 6.26E-04 | 0.12 |
| **Sum** | 22 | 0.11 | 2.55E-03 | <0.001 |

CC variants identified in MVP, listed by rsID. Count and frequency in MVPs are out of 203 MVPs tested, while frequency in general population is out of 730,947 exomes queried in gnomad v4.1.0. CC = cardiomyopathy/channelopathy; MVP = mitral valve prolapse.

## Abbreviations and Acronyms

AI: Artificial intelligence
CC: Cardiomyopathy and channelopathy
CI: Confidence interval
CMR: Cardiac magnetic resonance imaging
GLS: Left ventricular global longitudinal strain
HR: Hazard ratio
LA: Left atrium
LGE: Late gadolinium enhancement
LV: Left ventricle, left ventricular
LVEDVI: Left ventricular end-diastolic volume indexed
LVESVI: Left ventricular end-systolic volume indexed
MAD: Mitral annular disjunction
MD: Mechanical dispersion
MR: Mitral regurgitation
MVP: Mitral valve prolapse
OR: Odds ratio
P/LP: Pathogenic or likely pathogenic
SCA: sudden cardiac arrest
SCD: sudden cardiac death
STE: Speckle-tracking echocardiography
VAs: Ventricular arrhythmias

## Graphical Abstract

### CMR Acquisition and Image Analysis

CMR image acquisition was performed using 3.0T scanners Discovery MR750w or SIGNA Premier (GE Healthcare, Milwaukee, WI). Briefly, standard long- and short-axis cine images were obtained using a balanced steady-state free precession sequence. Typical parameters were TR/TE 2.8/1.3ms, flip angle 45°. Late gadolinium enhancement (LGE) imaging was performed 5-15 min following injection of 0.1 mmol/kg gadobutrol, and images were obtained using a high-resolution breath-hold two-dimensional sequence at three separate levels in the short-axis plane (basal, mid, and apical). An inversion-recovery fast gradient-echo sequence was performed in two phase-encoding directions to differentiate true late enhancement from artifact. The inversion time was optimized to achieve satisfactory nulling of the myocardium.

Image analysis was performed offline using a standardized approach (cvi42 version 5.14.0, Circle Cardiovascular Imaging Inc., Calgary, Canada). The presence of myocardial LGE was determined when part of the myocardial tissue present on the SSFP images was replaced by high signal intensity in the LGE images. Areas of inversion artefact or signal contamination by epicardial fat or blood pool were excluded.

